# An auditable multimodal agentic system for sequential decision-making across the acute stroke pathway

**DOI:** 10.64898/2026.09.24.26363786

**Authors:** Kai-cheng Yang, Zhijie Bao, Lu-Sha Tong, Jing Yuan, Yuxiao Chen, Xiaoyi Xue, Yueqi Zhu, Yunhe Luo, Jin Zheng, Xueyu Mao, Zhuohang Liu, Yong Wang, Zengyu Zhang, Rong Ji, Xuan Shi, Jiajie Peng, Zhongyu Wei, Jing Zhao

**Author notes:** Corresponding author(s). E-mail(s); zhao. These authors contributed equally to this work and share first authorship (Kai-cheng Yang, Zhijie Bao).

## Abstract

Ischemic stroke is a leading cause of long-term disability worldwide. Although timely reperfusion can improve outcomes, realizing this benefit requires clinicians to integrate fragmented clinical and imaging information with evolving evidence under the time pressure of emergency care, often with limited access to specialist expertise. Here we present StrokeAgent, a multimodal agentic decision-support framework designed around temporal alignment and continuous information synthesis across the acute stroke pathway. It processes clinical records, laboratory results, raw electrocardiographic (ECG) waveforms and source multimodal CT images to support emergency triage, the initial thrombolysis decision and final reperfusion planning. As information becomes available, StrokeAgent formulates and updates recommendations at each decision point, grounding each recommendation in an auditable chain of supporting findings and evidence. We evaluated the system in a curated set of 100 patients with multimodal clinical and imaging data, deliberately enriched for clinically complex reperfusion scenarios alongside standard presentations. StrokeAgent achieved 86% concordance with expert-adjudicated final decisions, exceeding the mean performance of three tool-free large language models (LLMs) and six resident and attending neurologists by 13 and 17 percentage points, respectively, under matched information content and timing. Complete-pathway concordance (agreement at every applicable decision point) was 76%, with even larger margins over both comparator groups. On average, StrokeAgent completed the full decision pathway 73.9 s faster than the case-level median neurologist time. In 1,200 blinded clinician assessments, StrokeAgent received higher overall clinical utility ratings than each LLM (common odds ratios, 10.8–16.3) and higher reasoning-quality ratings, with fewer outputs judged to pose severe potential harm. StrokeAgent demonstrates the feasibility of a decision-support system that integrates native multimodal data processing with auditable, evidence-supported recommendations aligned with the sequential nature of acute stroke care. Prospective studies are needed to determine whether physician-supervised use improves clinical care.

## 1 Main

Ischemic stroke remains a major contributor to death and disability worldwide, accounting for approximately 3.28 million deaths and 67 million disability-adjusted life years in 2023[1]. For appropriately selected patients, intravenous thrombolysis (IVT) and endovascular thrombectomy (EVT) improve functional outcomes, but realizing these benefits requires clinicians to integrate complex clinical and imaging information rapidly, as treatment benefit diminishes with delay[2–8]. Yet the specialist stroke expertise needed to navigate these time-critical decisions remains scarce, contributing to substantial disparities in the quality of acute stroke care at lower-tier hospitals and during off-hours. For example, only 42.2% of eligible patients in Chinese secondary hospitals received IVT under usual care[9], with off-hours admission associated with further treatment delays[10].

Acute stroke care is a highly constrained race against time conducted amid incomplete and evolving information[3]. Clinicians cannot wait for a complete diagnostic picture; they must formulate and update treatment strategies as heterogeneous data streams emerge sequentially[2, 11]. This principle is reflected in major clinical guidelines, which recommend initiating IVT in eligible patients without delaying treatment for additional vascular or perfusion imaging[2, 12, 13]. The core clinical difficulty, therefore, lies in making high-stakes decisions as partial information unfolds. While conventional deep-learning models have been extensively explored for stroke diagnosis and abnormality detection[14–17], these static paradigms are ill-suited to the temporal dynamics of decision-making.

Agentic artificial intelligence (AI) extends these capabilities by using large language models (LLMs) to coordinate tools and medical knowledge across multistep tasks[18–24]. Such systems have emerged in ophthalmology and oncology[20, 22], with recent work extending to treatment recommendations in stroke[24]. Yet several gaps persist between current agentic frameworks and the realities of acute stroke care. First, existing systems are rarely designed to be embedded in the continuous clinical workflow; they are typically evaluated on completed vignettes rather than on their ability to navigate the sequential revelation of clinical information. Second, they remain poorly aligned with the multimodal reality of emergency care, typically relying on prestructured text or preprocessed image representations rather than working directly from physiological signals and source neuroimaging. Third, few systems are designed to keep high-stakes recommendations transparent and independently verifiable as the decision chain evolves. What is needed is a decision-support system aligned with the demands of acute stroke care—one that can work directly from the heterogeneous data acquired during emergency care, adapt its recommendations as the clinical picture evolves, and expose an auditable evidentiary trail for each recommendation.

Here we present StrokeAgent, a multimodal agentic framework built for the dynamic, staged course of acute stroke care, with a design philosophy centered on temporal alignment and continuous information synthesis. The architecture natively integrates specialized tools and clinical subagents capable of processing routine emergency-care data streams ranging from unstructured electronic health records and raw ECG waveforms to source multimodal CT imaging. As the emergency work-up progresses, the system continuously assimilates this newly available multimodal information, seamlessly navigating the care trajectory from emergency triage, through initial thrombolysis decision, to final reperfusion planning. The processed findings are then linked to a curated evidence base spanning 30 reperfusion trials and more than 100 guideline-based rules developed and refined by experts, to inform treatment recommendations. Each recommendation remains traceable throughout the pathway, accompanied by intermediate findings, tool-derived imaging outputs and citations to the evidence that supports it, enabling clinician review.

We evaluated StrokeAgent on a curated set of 100 suspected acute stroke cases to capture the complexity of real-world reperfusion scenarios, with deliberate enrichment for extended or uncertain time windows, relative contraindications and large-core infarction, alongside hemorrhage and stroke mimics. To establish a robust benchmark, five domain experts adjudicated the reference standards, and the system’s performance was evaluated against both tool-free LLMs and a panel of six resident and attending neurologists. Crucially, all evaluations were conducted under the same staged information boundaries to ensure a fair comparison of sequential decision-making capabilities. StrokeAgent achieved 86% concordance with expert final decisions and 76% complete-pathway concordance (agreement at every applicable decision point within a case), exceeding the corresponding group means by 13.0 and 29.0 percentage points for the tool-free LLMs and by 17.0 and 22.2 percentage points for the neurologists. StrokeAgent completed the full decision pathway a mean of 73.9 s faster than the case-level median neurologist time. We further assessed the clinical utility and quality of the reasoning supporting these decisions through blinded review of the 400 trajectories produced by StrokeAgent and the three LLMs. StrokeAgent received higher overall clinical utility ratings than each LLM and higher ratings on all four prespecified quality dimensions: factual correctness, completeness, stage appropriateness and reasoning–decision consistency. A lower proportion of its outputs were judged to pose severe potential harm (2% versus 8–16% for the LLMs). Full implementation and evaluation procedures are provided in Methods, with an overview of the pipeline in Fig. 1.

**Fig. 1.**
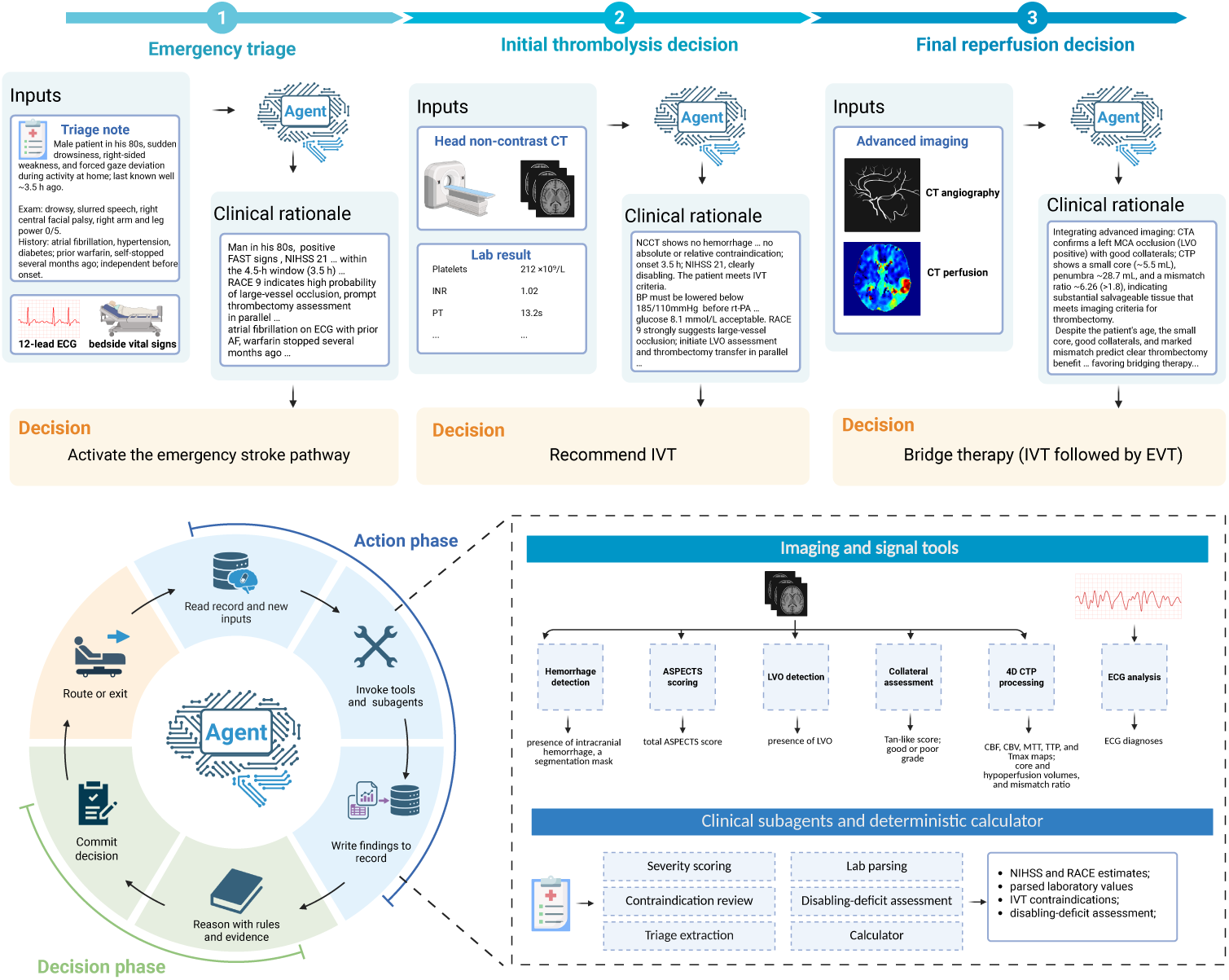
Overview of StrokeAgent. StrokeAgent processes clinical and imaging information directly from source data as it becomes available through three stage-specific agents for emergency triage, initial thrombolysis decision and final reperfusion planning. At each stage, specialized tools and clinical subagents return structured findings to a shared case record, which the agent reasons over together with guideline-based rules and matched trial evidence to formulate a stage-specific recommendation. The upper panel shows an illustrative case progressing through the three decision stages. The lower panels summarize the agent decision cycle and the processing components available to the system. Illustration created with BioRender.com.

## 2 Results

### 2.1 StrokeAgent achieves expert-concordant decisions across the staged workflow

We first assessed how well StrokeAgent’s decisions agreed with the expert reference across the 100 evaluation cases. The primary endpoint was final clinical decision concordance, defined as exact agreement on the final action, selected from six mutually exclusive options. After adjudication, the expert reference comprised EVT alone in 36 cases, bridging therapy in 18, IVT alone in 18, conservative management in 18, hemorrhage in 5 and stroke-mimic in 5. StrokeAgent matched the expert reference in 86 of 100 cases (86.0%, 95% CI 79.0–92.0%; Cohen’s *κ* = 0.817; Fig. 2a).

**Fig. 2.**
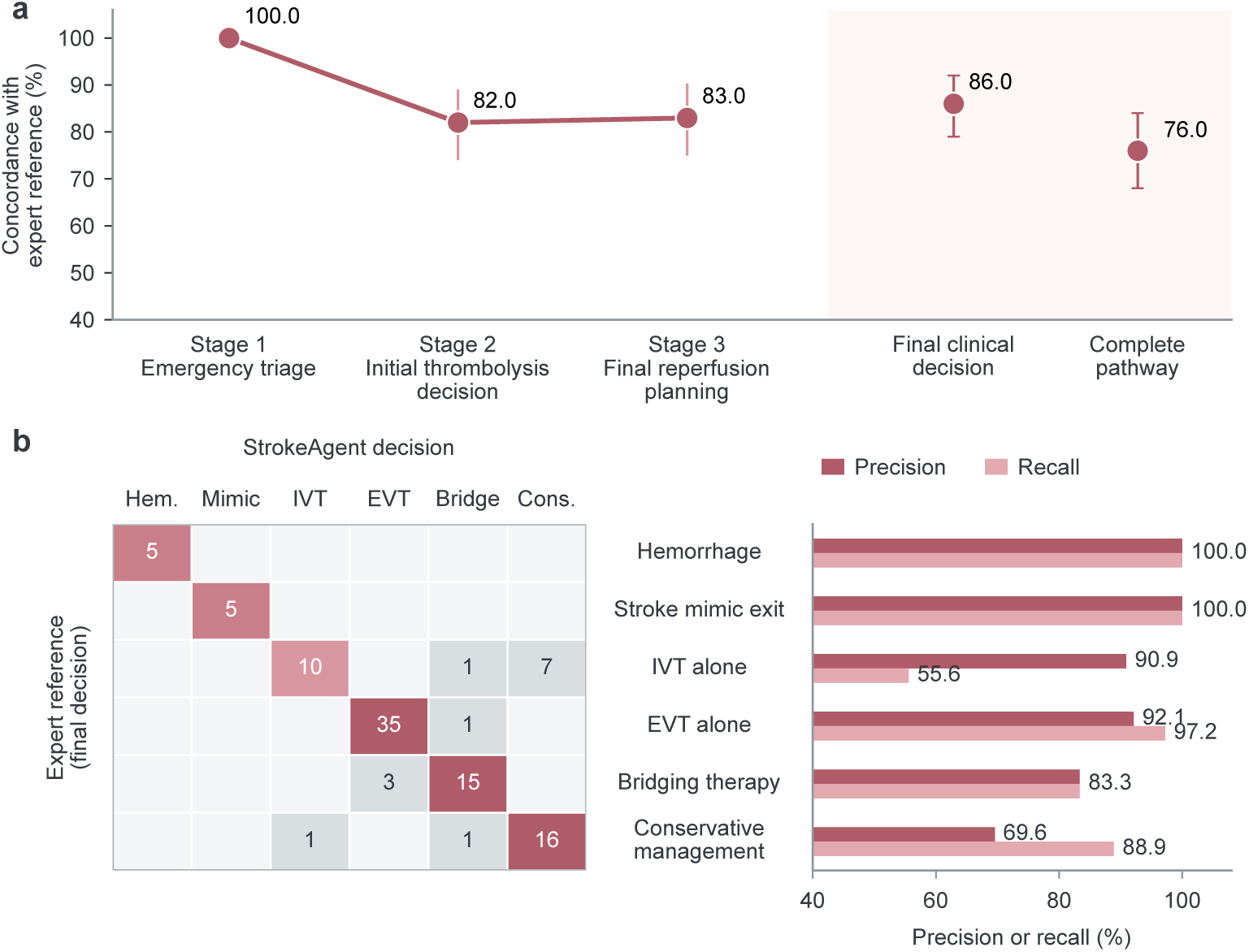
StrokeAgent performance across the acute stroke pathway. a, Concordance between StrokeAgent and the expert reference across the three sequential decision points, together with final clinical decision concordance and complete-pathway concordance. Complete pathway concordance required agreement with the expert reference at every applicable decision stage within a case. Points indicate proportions and error bars indicate 95% confidence intervals. b, Confusion matrix comparing StrokeAgent decisions with the expert reference across the six mutually exclusive final clinical actions. Numbers within cells indicate case counts. Horizontal bars show class-specific precision and recall (%) Hem., hemorrhage exit; Mimic, stroke-mimic exit; IVT, intravenous thrombolysis alone; EVT, endovascular thrombectomy alone; Bridge, bridging intravenous thrombolysis and endovascular thrombectomy; Cons., conservative management.

A correct final decision can nevertheless follow an incorrect intermediate one. We therefore also assessed complete-pathway concordance, which required agreement at every applicable decision point. StrokeAgent met this stricter criterion in 76.0% of cases. Stage-level concordance was 100.0% at triage, 82.0% at the initial thrombolysis decision and 83.0% at final reperfusion planning.

At the final-action level, the macro-averaged F1 was 0.875 (Fig. 2b). Performance was highest for hemorrhage and stroke-mimic exits, both with 100% precision and recall, whereas IVT alone had the lowest recall (10 of 18 cases, 55.6%) despite a precision of 90.9%. Conservative management showed lower precision (69.6%), reflecting cases in which the expert reference indicated reperfusion, but StrokeAgent recommended conservative management.

### 2.2 StrokeAgent outperforms tool-free LLMs and neurologists

We next benchmarked StrokeAgent against three LLM baselines (GPT-5.4, Gemini 3.1 Pro and Claude Opus 4.6; hereafter GPT-5.4, Gemini and Claude) that had no access to StrokeAgent’s processing components or evidence layer. At each stage, the baselines received the same clinical information and imaging modalities as StrokeAgent, in the same temporal sequence. However, because the LLMs could not ingest DICOM series directly, the series were converted into standard image formats for them, whereas StrokeAgent operated on the source DICOM series throughout. To place StrokeAgent’s performance in clinical context, we also compared it with six neurologists (three residents and three attendings) from five comprehensive stroke centers. They evaluated the same 100 cases under the same staged information boundaries using only traditional sources. Before committing to a decision, they also had to derive the intermediate findings themselves from the available modalities, for example by recognizing large vessel occlusion (LVO).

StrokeAgent had higher final clinical decision concordance than both comparator groups. Concordance was 86.0% for StrokeAgent and averaged 73.0% across the three LLMs, corresponding to an absolute difference of 13.0 percentage points (95% CI 5.7–20.7; Holm-adjusted P = 6.8 *×* 10*^−^*^4^; Fig. 3a, b). Individual LLM performance ranged from 71.0% to 75.0%, with StrokeAgent outperforming each individually (all Holm-adjusted P *<* 0.02, case-level results in Fig. 3c). The mean final clinical decision concordance across the six neurologists was 69.0%, with individual performance spanning 62.0–76.0%; StrokeAgent exceeded this mean by 17.0 percentage points (95% CI 9.5–24.3; Holm-adjusted P = 1.7 *×* 10*^−^*^5^).

**Fig. 3.**
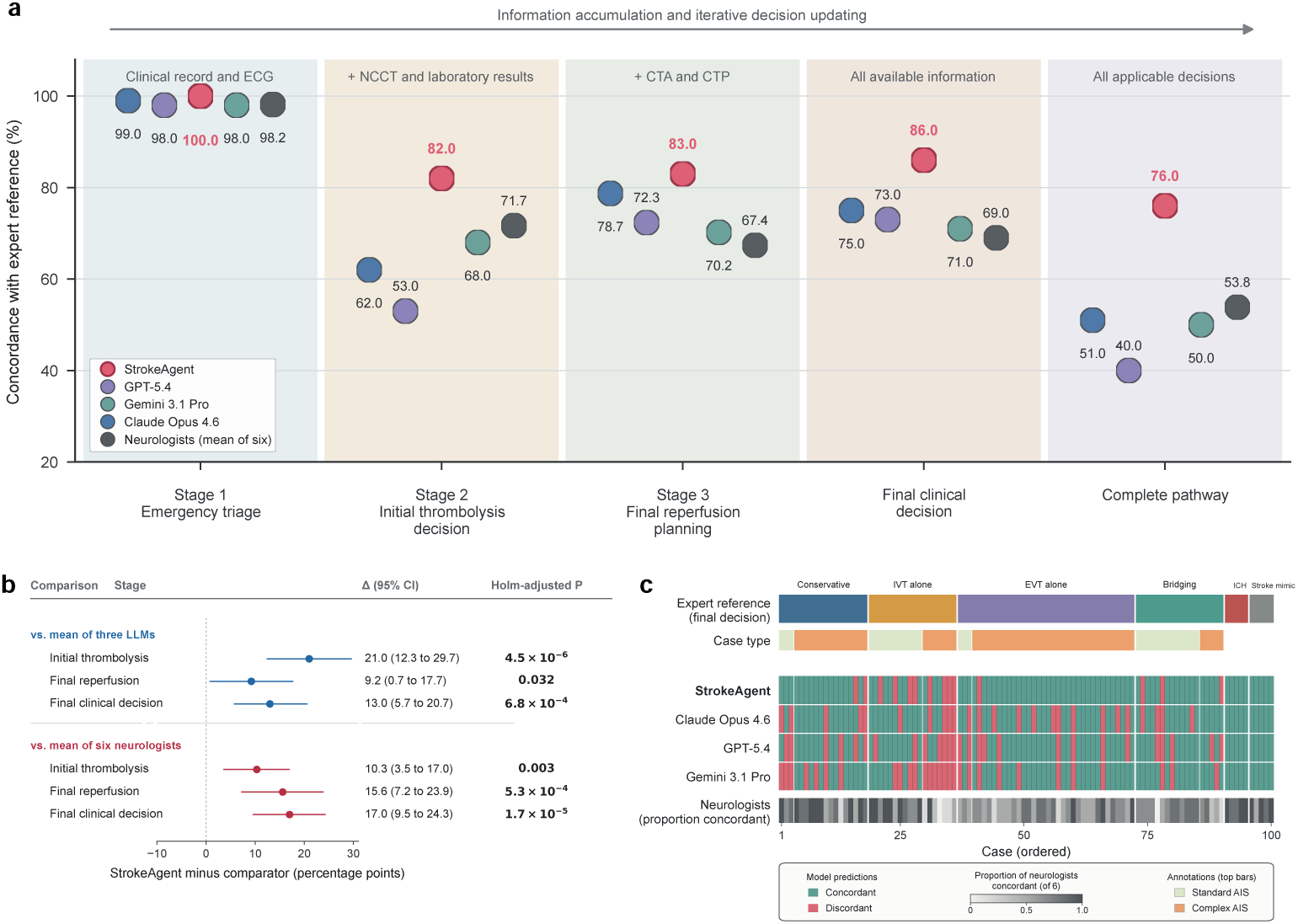
Comparative performance of StrokeAgent, tool-free LLMs and neurologists. a, Concordance with the expert reference for StrokeAgent, three tool-free LLMs and six neurologists at each decision point, for the final clinical decision and across the complete pathway. Complete-pathway concordance is defined as in Fig. 2a. b, Differences in concordance between StrokeAgent and the mean of the three LLMs and of the six neurologists. Points indicate percentage-point differences and horizontal lines indicate 95% confidence intervals (10,000 case-level bootstrap resamples); P values are Holm-adjusted. c, Case-level final-decision concordance across the 100 evaluation cases. Shading for neurologists indicates the proportion of the six neurologists whose final decisions were concordant with the expert reference. Cases are ordered by the expert reference final decision; the upper annotation bars indicate the reference action and whether the case was standard or complex.

The difference was greater when agreement was required throughout the decision pathway. Complete-pathway concordance was 76.0% for StrokeAgent, compared with 40.0–51.0% for the LLMs (mean 47.0%) and 49.0–61.0% for the neurologists (mean 53.8%). Relative to the respective group means, the advantage increased from 13.0 percentage points at the final decision to 29.0 percentage points across the complete pathway for the LLMs (95% CI, 20.3–37.7; Holm-adjusted P = 1.3 *×* 10*^−^*^10^), and from 17.0 to 22.2 percentage points for the neurologists (95% CI, 13.8–30.3; Holm-adjusted P = 8.8 *×* 10*^−^*^8^). The largest between-group differences occurred at the two treatment decisions. Triage concordance was near ceiling for all groups (98.0–100.0%). However, StrokeAgent reached 82.0% concordance at the initial thrombolysis decision, compared with 53.0–68.0% for the LLMs and 68.0–75.0% for the neurologists. Across the three LLMs, IVT was recommended in 49 of 93 instances in which the reference decision was IVT (52.7%), whereas 34 were instead deferred for further evaluation. At final reperfusion planning, concordance was 83.0% for StrokeAgent, compared with 70.2– 78.7% for the LLMs and 59.6–74.5% for the neurologists (n = 94 applicable cases). Of the 184 neurologist errors at this stage, 65 (35.3%) involved discordance between EVT alone and bridging therapy.

Across the 100 cases, median end-to-end decision time was 258.1 s (IQR, 235.8– 282.2) for StrokeAgent and 315.7 s (IQR, 271.8–382.9) for the case-level median neurologist time. The mean paired difference (StrokeAgent minus neurologists) was -73.9 s (bootstrap 95% CI, -89.2 to -59.1; P = 8.5 *×* 10*^−^*^16^). StrokeAgent required less median time for triage (30.6 versus 64.8 s) and the initial thrombolysis decision (55.2 versus 81.0 s), but more time for final reperfusion planning (172.0 versus 156.6 s), a stage at which StrokeAgent derived perfusion outputs from source computed tomography perfusion (CTP) data whereas neurologists received them precomputed.

### 2.3 Blinded clinicians rate StrokeAgent’s decision trajectories as more clinically useful and better reasoned

Six blinded neurologists rated the clinical rationales accompanying each decision trajectory, with each of the 400 outputs (100 cases *×* 4 systems) scored independently by three raters, yielding 1,200 ratings. For overall clinical utility, StrokeAgent was rated higher than each tool-free LLM (Fig. 4a): compared with GPT-5.4, the common odds ratio (OR) was 10.78 (95% CI, 4.76–24.44; Holm-adjusted P = 1.2 *×* 10*^−^*^8^, Fig. 4b); compared with Gemini, 11.52 (5.07–26.17; Holm-adjusted P = 1.1 *×* 10*^−^*^8^); and compared with Claude, 16.29 (7.12–37.28; Holm-adjusted P = 1.2 *×* 10*^−^*^10^). At the case level, an output was considered favorable when its median utility rating across the three raters was 4 or 5. StrokeAgent produced favorable outputs in 84 of 100 cases, compared with 65, 63 and 62 for GPT-5.4, Gemini and Claude (Fig. 4c); the pairwise differences ranged from 19 to 22 percentage points (Holm-adjusted P = 3.4 *×* 10*^−^*^4^ to 1.9 *×* 10*^−^*^3^).

**Fig. 4.**
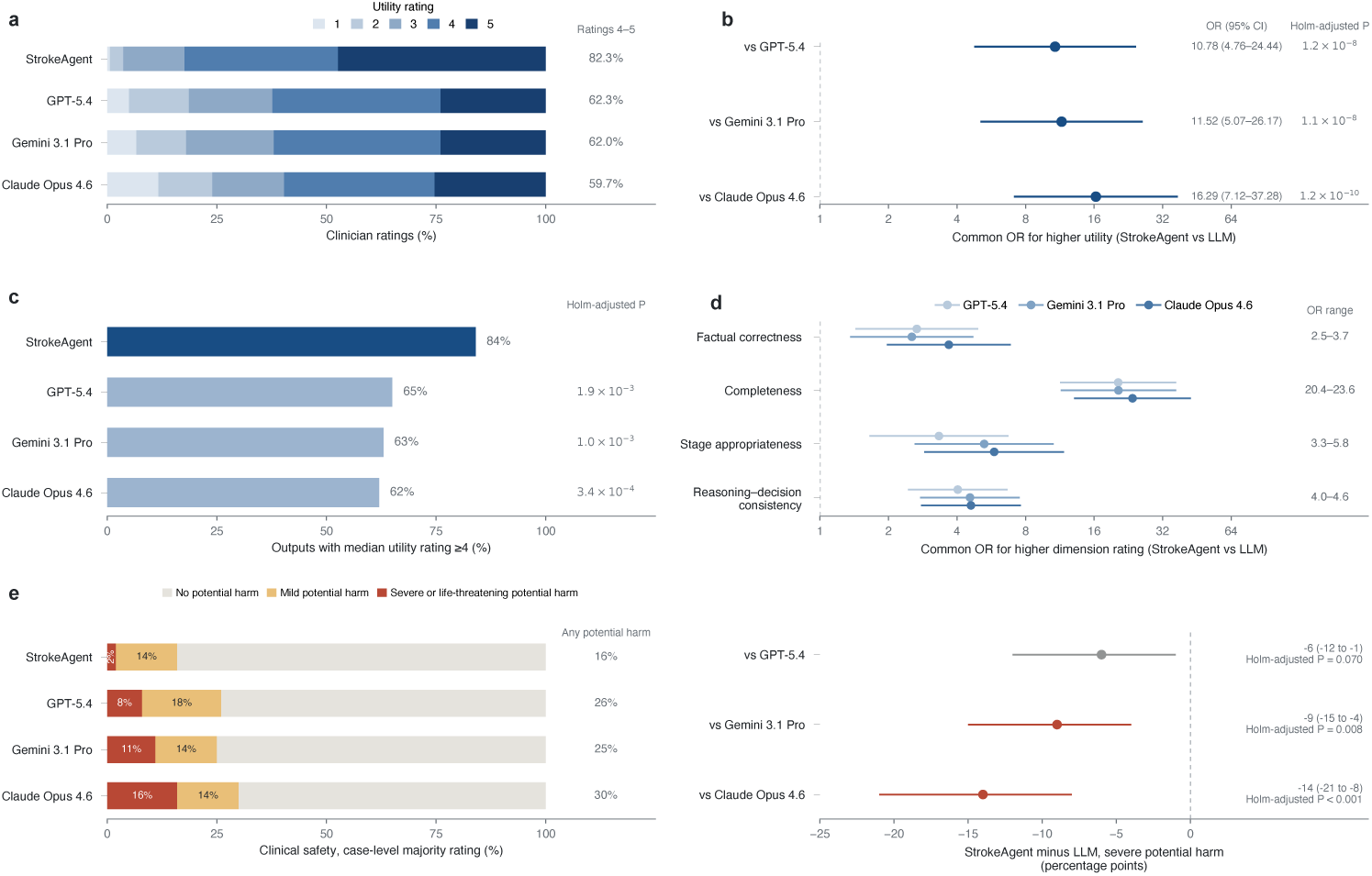
Blinded clinician evaluation of StrokeAgent and LLM rationales. Six neurologists rated 400 outputs (100 cases *×* 4 systems), each rated by three raters (1,200 ratings; 300 per system). a, Distribution of overall clinical utility ratings for StrokeAgent and three tool-free LLMs. b, Common ORs for higher clinical utility ratings for StrokeAgent versus each LLM. c, Proportion of outputs with a median clinical utility rating of 4 or 5 across the three raters. d, Common ORs for higher ratings across the four secondary quality dimensions. e, Left, clinical safety at the case level (majority rating of the three raters); right, differences in severe potential harm between StrokeAgent and each LLM. Points and horizontal lines in b, d and e indicate effect estimates and 95% confidence intervals.

Across the four secondary prespecified quality dimensions (factual correctness, completeness, stage appropriateness and reasoning–decision consistency), all 12 comparisons favored StrokeAgent after multiplicity adjustment (P *≤* 4.0 *×* 10*^−^*^3^, Fig. 4d). The largest differences were observed for completeness (OR range, 20.4–23.6), for which 95.7% of StrokeAgent ratings were 4 or 5 compared with 59.3–60.0% for the LLMs. Differences were also observed for stage appropriateness (OR range, 3.3–5.8), factual correctness and reasoning–decision consistency (OR range, 2.5–4.6).

Safety was analyzed as the case-level majority rating. Outputs were judged to pose severe potential harm in 2% of cases for StrokeAgent, compared with 8% for GPT-5.4, 11% for Gemini and 16% for Claude (Fig. 4e). StrokeAgent outputs were less often judged to pose severe potential harm than those of Gemini (difference, - 9 percentage points; Holm-adjusted P = 0.008) and Claude (-14 percentage points; Holm-adjusted P *<* 0.001), whereas the difference from GPT-5.4 was not statistically significant (-6 percentage points; Holm-adjusted P = 0.070). StrokeAgent also had the lowest frequency of any potential harm (16% versus 25–30% for the LLMs), although the overall difference was not significant (Holm-adjusted P = 0.055).

Inter-rater agreement (ordinal Krippendorff’s *α*) was 0.669 for overall clinical utility, 0.454 to 0.524 across the four quality dimensions and 0.824 for clinical safety.

### 2.4 Traceable outputs support error analysis

We reviewed all 14 cases in which StrokeAgent’s final clinical decision differed from the expert reference. For each case, we examined the source clinical record, component outputs and complete decision trajectory, and assigned the primary error mechanism by consensus (Fig. 5). Seven were attributed to clinically meaningful errors in tool or subagent outputs and seven to errors in decision synthesis without any material factual error. Component errors comprised incorrect National Institutes of Health Stroke Scale (NIHSS) or disabling-deficit assessments (n = 3), LVO calls (n = 2), contraindication screening (n = 1) and an Alberta Stroke Program Early CT Score (ASPECTS) estimate (n = 1). These errors typically propagated downstream, leaving subsequent decisions internally coherent but based on a false premise. Whereas component errors mostly arose by the initial thrombolysis decision (5 of 7), synthesis errors occurred more often during reperfusion planning (5 of 7) and reflected unsupported inference (n = 3) or inappropriate weighting of treatment benefit and risk (n = 4). For example, StrokeAgent treated conditions that are not guideline-defined contraindications, such as myocardial infarction 2 years earlier or chronic kidney disease with renal anemia, as additional bleeding concerns and withheld IVT that the expert had recommended. By contrast, in one case with prior clipping of a ruptured intracranial aneurysm, StrokeAgent recommended IVT, the more aggressive option, whereas the expert reference favored conservative management. Ten of the 14 discordances involved omission or de-escalation of reperfusion therapy recommended by the expert reference, whereas four involved treatment escalation beyond the reference.

**Fig. 5.**
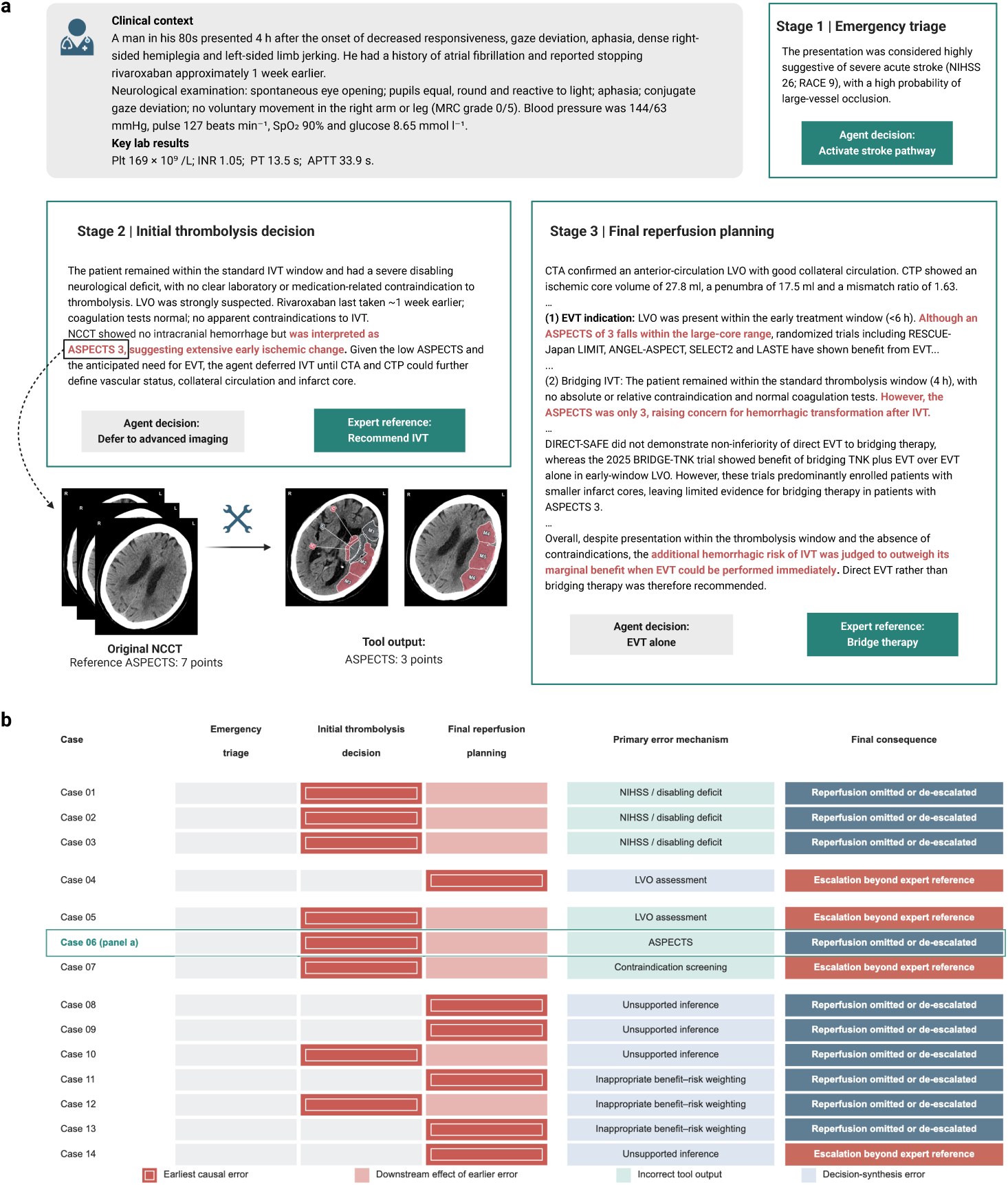
Expert audit of StrokeAgent decision trajectories and error localization. a, Representative audit of a discordant decision trajectory (case 06), tracing an incorrect ASPECTS estimate from the initial thrombolysis decision to final reperfusion planning. Red text marks the erroneous tool-derived content and its downstream use; rationale text is abridged from the StrokeAgent output. Reference ASPECTS is the expert-rated value 7 b, Case-level localization of the earliest causal error (outlined) and its downstream effect (shaded) across the 14 discordant final decisions, with the primary error mechanism (incorrect tool output or decision-synthesis error) and the final consequence relative to the expert reference. Case 06 corresponds to panel a.

**Fig. 6.**
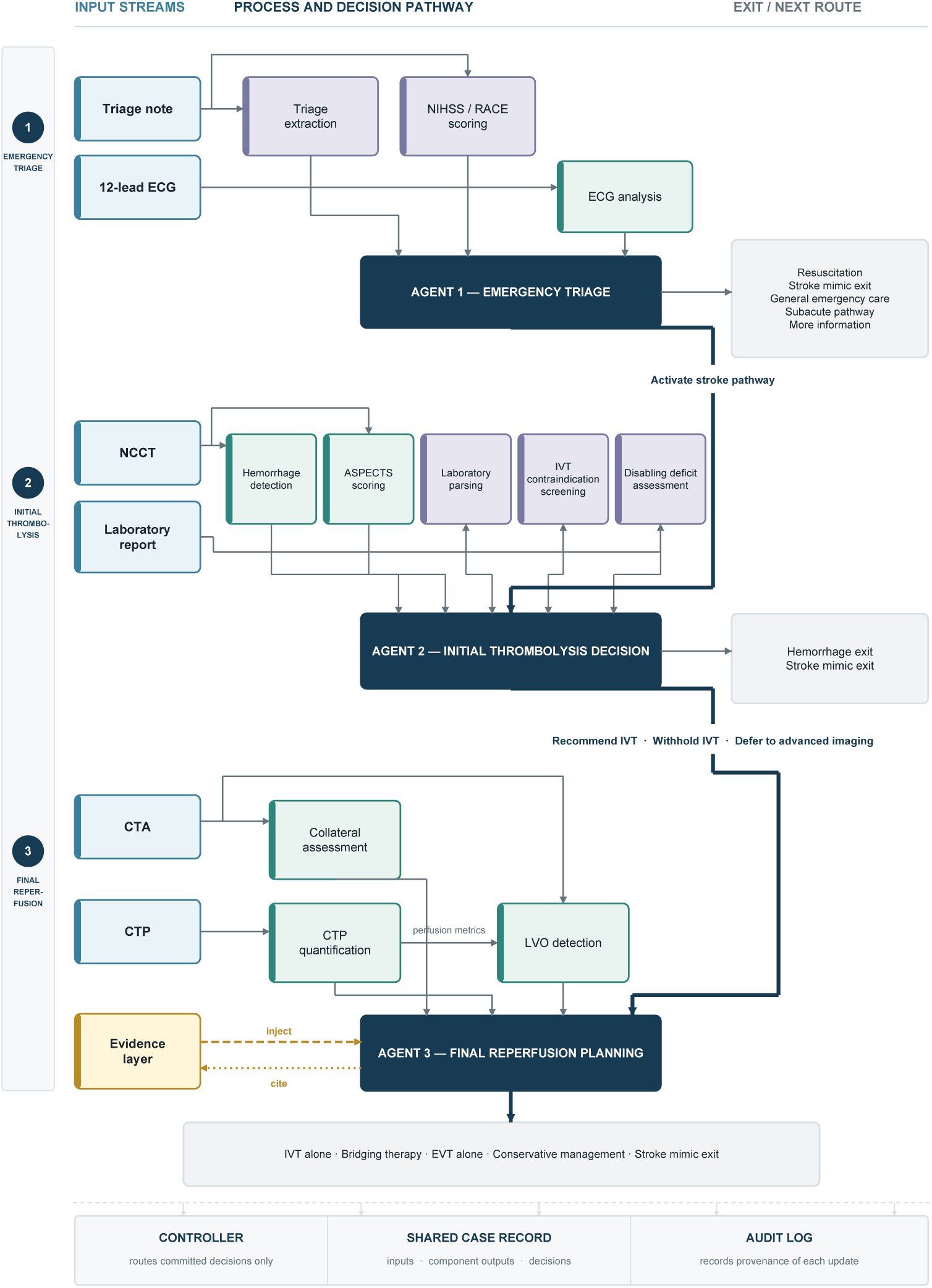
Staged multimodal decision architecture of StrokeAgent. StrokeAgent processes sequentially available clinical, ECG, laboratory and imaging information through three decision agents: emergency triage, initial thrombolysis decision, and final reperfusion planning. Each agent integrates inputs with task-specific components and produces either a terminal exit or a committed decision that is passed to the next stage. The trial evidence base is supplied to the reperfusion agent as context and cited in its output. All component outputs and agent decisions are written to a shared case record, while the controller routes committed decisions and the audit log records the provenance of each update. Blue boxes indicate input streams, green boxes indicate specialized tools, purple boxes indicate LLM-based clinical subagents, dark-blue boxes indicate decision agents, yellow indicates the evidence layer, and grey boxes indicate terminal outputs and shared system infrastructure.

## 3 Discussion

We developed StrokeAgent, a multimodal agentic system designed to support decision-making across the acute stroke care pathway, from emergency triage to reperfusion planning, and conducted an initial evaluation on a carefully curated case set. Our work makes three main contributions. First, StrokeAgent provides end-to-end decision support through temporal alignment with the stroke care pathway and continuous integration of evolving information. Second, we designed a multidimensional evaluation framework that more closely reflects the conditions under which clinical decisions are made. Third, StrokeAgent demonstrated advantages across the entire decision trajectory, in the quality of the supporting rational, and in overall decision speed.

StrokeAgent was designed to align automated multimodal data processing with the temporal structure of decision-making in acute stroke care. In practice, clinical information is fragmented, time-dependent and distributed across systems, increasing clinicians’ cognitive burden of information integration[2, 25]. StrokeAgent addresses this challenge by directly processing the heterogeneous source data required for decision-making within a unified workflow, without manual data organization or transcription and without reliance on preprocessed imaging measurements or prestructured inputs. For example, CTP can be processed directly for perfusion analysis and incorporated into subsequent decision-making, reducing the need for cross-system data transfer and intermediate processing. This architecture is intended to reduce the operational burden of information handling and free clinicians to focus on patient care. StrokeAgent also supports stage-aware dynamic decision-making, formulating recommendations based on the information available at each decision point and updating them as new information emerges. For instance, once hemorrhage and contraindications have been excluded, StrokeAgent can recommend IVT for an eligible patient; if subsequent CT angiography (CTA) identifies LVO, the recommendation can be updated to bridging therapy. Recent agentic systems have begun to move beyond static, single-pass clinical reasoning, and our work extends this direction from solving a bounded clinical task to participating continuously in decision-making along the clinical timeline[23, 26]. Finally, evolving guidelines and clinical evidence make it an ongoing challenge to stay current. StrokeAgent links clinical findings to guideline-based rules and relevant evidence, so that treatment recommendations are explicitly constrained by the supporting evidence, while its knowledge and evidence layers can be updated as guidelines and trial evidence evolve.

As clinical AI systems become increasingly interactive and agentic, conventional evaluations based on static case descriptions or a single final answer are no longer sufficient to capture their performance in real-world decision-making[26–28]. Although recent benchmarks have begun to address this limitation through sequential information acquisition and simulated clinical workflows[23, 26], in acute stroke, evaluating only the final recommendation may obscure whether earlier decisions were appropriate when they had to be made. We therefore evaluated StrokeAgent and the comparator systems under the same staged information boundaries and operating procedures. The expert reference was constructed under the same information constraints: at each stage, experts determined the preferred management strategy using only the information available at that time, the 2026 American Heart Association/American Stroke

Association (AHA/ASA) guideline evidence[2], and their clinical expertise, rather than using the care actually delivered as the reference. This distinction matters because delivered treatment reflects not only clinical indications but also patient or family preferences and institutional practice patterns, and is therefore an imperfect benchmark of decision quality. In addition, we carefully curated the case set to include challenging reperfusion scenarios that demand complex imaging interpretation and evidence weighing, allowing us to assess StrokeAgent’s performance across diverse clinical conditions. The set also varied in imaging availability, including 15 cases without CTP, which allowed assessment under resource-constrained conditions.

Within this evaluation framework, StrokeAgent’s advantage was more pronounced across the complete pathway than at the final decision alone. This distinction is clinically important because an inappropriate decision in an early treatment window may not be fully recoverable even if corrected later, since time lost cannot be regained in the race to salvage brain tissue. In particular, tool-free LLMs frequently deferred IVT in cases where the expert panel considered the available information sufficient to support treatment at that time. StrokeAgent also showed a speed advantage, concentrated in the earlier stages of the pathway. At final reperfusion planning it took longer than neurologists, who were given precomputed CTP perfusion maps and volumetric parameters, whereas StrokeAgent’s time included generating these outputs from source CTP data; the overall speed advantage was thus achieved despite this additional step. The system’s advantage also extended to the quality of the reasoning supporting its decisions. For clinical decision-support systems, final-decision accuracy alone is insufficient; the factual correctness, completeness and clinical utility of the rationale also determine whether a recommendation can be meaningfully reviewed and adopted[28, 29]. StrokeAgent’s traceable architecture further enabled error localization, distinguishing upstream component errors from downstream decision-synthesis errors. Beyond allowing clinicians to verify or challenge a recommendation at the point of care, this distinction clarifies priorities for refining the system: the former requires better perception or information extraction, whereas the latter requires improved integration and weighting of otherwise accurate findings. Taken together, these results suggest that the value of clinically oriented agentic systems lies not only in sustaining accuracy across sequential decisions, but also in the quality of the clinical rationale supporting those decisions and in overall decision speed.

Looking ahead, StrokeAgent may be most valuable as a scalable decision-support layer within stroke systems of care, particularly in hospitals with limited on-site neurological expertise. Rural and resource-limited centers continue to face gaps in access to timely reperfusion care[30], and AI-enabled virtual stroke networks are increasingly being explored as a means of extending specialist capability across institutions[31]. By aligning with existing stroke-care workflows and operating directly on routinely acquired source data, StrokeAgent may be readily integrated into clinical practice across different care settings. Its modular architecture supports such adaptation: its processing components and evidence layer can be maintained and updated independently as local infrastructure, clinical pathways and treatment evidence evolve. Rather than replacing stroke specialists, StrokeAgent could offer an updatable, auditable second opinion at the point of care.

Our study has several limitations. First, StrokeAgent follows a three-stage structure with predefined steps at each stage. Although this captures the major decision points in the reperfusion pathway, real-world stroke care is less linear than this structure implies: imaging, transfer and treatment preparation may proceed in parallel, and workflows vary with local resources and logistics. The present architecture should therefore be viewed as an abstraction of the principal reperfusion pathway rather than a reproduction of the full complexity of bedside care. Second, StrokeAgent’s performance is bounded by the accuracy of its processing components. Several of these were adapted from open-source methods, with fine-tuning or calibration on limited datasets. We provide tool-level performance in the Extended Data Fig. 1 and Supplementary Tables 1–3, but these results represent a preliminary evaluation under our study conditions, and upstream errors can influence downstream decisions. The modular architecture, however, allows individual components to be replaced with better-validated tools as they become available, which may mitigate this risk in future iterations.

Third, the evaluation set was relatively small, retrospectively collected from three hospitals in eastern China and a public dataset; the case mix, which was deliberately enriched for complex scenarios, does not reflect the distribution seen in an unselected emergency stroke population. For the 25 cases drawn from the Ischemic Stroke Lesion Segmentation (ISLES) 2024 challenge dataset, imaging came from real patients, but the clinical records were constructed from the limited information provided and may be more complete than routine emergency documentation. The neurologist group comprised only six clinicians, spanning a limited range of experience. All groups received information in the same sequence, but the format of imaging inputs differed. In particular, the comparator groups received post-processed perfusion maps and volumetric measurements whereas StrokeAgent generated these outputs itself from source image data, which also affected the time comparison at final reperfusion planning. We also did not test physicians working with StrokeAgent and therefore cannot determine whether the system improves physician performance. Accordingly, the present study should be viewed as an initial evaluation of agentic decision support across the acute stroke pathway. Future work will prospectively evaluate physician–StrokeAgent collaboration and system performance under real-world deployment conditions.

## 4 Methods

### 4.1 StrokeAgent architecture

StrokeAgent receives multimodal data acquired during the emergency stroke evaluation, including free-text clinical records, laboratory results, bedside measurements, 12-lead ECG waveforms, and source non-contrast CT (NCCT), CTA and 4D CTP acquisitions. Three decision agents are arranged in sequence, each corresponding to a decision point in the emergency stroke pathway: an emergency triage agent, an initial thrombolysis decision agent, and a final reperfusion planning agent. They share a common reasoning pattern and operate on a continuously updated shared case record. Supporting the decision agents is a processing layer with two classes of callable components: specialized tools for image, signal and quantitative processing, and clinical subagents for higher-order assessments that require synthesis of free-text clinical information. A rule and evidence layer provides more than 100 stage-specific, guideline-based rules, together with structured summaries of 30 primary reperfusion trials. The shared case record retains all inputs, component outputs and decisions, and an audit log records their provenance. The three decision agents were powered by Claude Opus 4.6 (model specifications in Supplementary Information).

Each agent operates through two decoupled phases. In the action phase, which mirrors how a clinician gathers information before committing to a decision, it reads the shared case record and newly available inputs, invokes the relevant tools or clinical subagents and writes the structured findings they return to the record. In the decision phase, it reasons over the accumulated findings, applicable clinical rules and any matched trial evidence to select one of a predefined set of actions and route the case.

### 4.2 Three-stage workflow aligned with the emergency stroke timeline

Inputs are introduced in the order in which they become available during the emergency stroke work-up. The triage agent acts on arrival at the emergency department, before any imaging or laboratory results are available. It receives the emergency admission record, bedside measurements and 12-lead ECG waveforms. From these inputs it derives structured findings, including presenting symptoms, the reperfusion time window, medical and medication history, stroke severity and ECG interpretation. The decision phase then commits to one of the predefined triage actions, including activation of the stroke pathway, a stroke mimic exit, or routing to general emergency care (for non-stroke conditions or cases outside the 24-h time window).

The initial thrombolysis decision agent takes over once the first head NCCT is available. At this stage the agent establishes whether hemorrhage is present, whether marked early ischemic change is already visible, whether laboratory results and history indicate contraindications to IVT, and whether the current neurological deficit is disabling. The decision outputs at this stage are hemorrhage exit, stroke mimic exit, recommend IVT, withhold IVT, and defer to advanced imaging.

The final reperfusion planning agent adds findings obtained from vascular and perfusion imaging to the shared case record. Reasoning over the complete multimodal picture, it weighs guideline rules against the matched trial evidence and commits to one of five terminal outputs: IVT alone, bridging therapy (IVT followed by endovascular thrombectomy), EVT alone, conservative management, or stroke mimic exit.

The action sets of the first two agents contain both hand-off targets, which pass control to the next agent, and exit targets, so the workflow forms a branching clinical pathway.

### 4.3 Tools and clinical subagents

To align the processing layer with routine hospital data flow, the imaging pipelines process source DICOM studies directly, with preprocessing and quantitative analysis performed within the system and no manual conversion into model-specific image formats. On NCCT, hemorrhage and early ischemic change were assessed by separate pipelines. For hemorrhage detection, we fine-tuned a published VIOLA-U-Net hemorrhage-segmentation model on 836 additional NCCT scans to reduce false-positive hemorrhage calls in the acute stroke setting while preserving sensitivity[32–34]. The fine-tuned model determines hemorrhage status and generates a segmentation mask. ASPECTS is calculated using an atlas-based pipeline. The basal ganglia and supraganglionic levels are first localized through registration to a standard CT template, after which an age-specific ASPECTS atlas is aligned with the corresponding patient images to transfer the ten regional labels[35]. Each region was then scored using prespecified bilateral attenuation differences, yielding the total ASPECTS[36]. The scoring parameters were optimized on 166 NCCT scans from a separate tuning cohort. For CTP, we used PyPeT to process source 4D CTP image and generate perfusion parameter maps[37], from which hypoperfused-tissue volume, ischemic core volume and mismatch ratio are derived. Because PyPeT’s specific deconvolution and post-processing procedures make conventional thresholds inapplicable, we recalibrated the core and hypoperfusion thresholds on 168 scans from a separate tuning cohort. LVO detection and collateral assessment are performed on CTA images. LVO assessment combines repeated multimodal-LLM interpretation with a whole-brain multiclass cerebrovascular segmentation model. The multimodal-LLM assessment uses CTA maximum-intensity-projection (MIP) renderings and axial slices, together with available NCCT and CTP renderings, quantitative perfusion measurements and clinical context. The segmentation model provides bilateral vessel-volume and continuity measures that serve as an orthogonal check in prespecified cases[38]. When an occlusion is present, the agent calls the collateral assessment tool, which segments the CTA arteries with an nnU-Net-based model[39], registers an atlas of bilateral middle cerebral artery (MCA) territories to the patient’s CTA space[40], and thresholds the ipsilateral-to-contralateral arterial volume ratio to report good or poor collateral status and a Tan-like score. The nnU-Net-based model used released weights, and the classification thresholds were fixed on 99 CTA scans. The ECG tool is a deep-learning model for abnormal ECG detection that processes 12-lead waveforms in HDF5 format[41], and outputs structured diagnoses such as atrial fibrillation, conduction block, and sinus bradycardia. The model was developed on 2,322,513 ECG examinations and used here without fine-tuning.

Five LLM-based clinical subagents perform the higher-order text assessments: one extracts structured fields from the free-text clinical record; one estimates NIHSS and Rapid Arterial Occlusion Evaluation (RACE) scores from symptom and examination descriptions[42]; one determines whether the presenting deficit is disabling; one extracts thrombolysis-relevant laboratory values and flags abnormalities; and one screens the medical history, medication exposure, and laboratory results for IVT contraindications[2]. Thrombolytic dosing is computed by a deterministic function implemented within the system, without LLM involvement.

Tools and clinical subagents are invoked autonomously by StrokeAgent. Each processing component is registered with a structured specification defining its function, required inputs, output fields and accepted data formats. Further details on tool development, tuning datasets and validation are provided in the Supplementary Tables 1-3; none of the tuning sets overlapped with the evaluation set.

### 4.4 Structured evidence library

More than 100 stage-specific clinical rules were authored and iteratively refined by a separate stroke-expert panel, based on the 2026 AHA/ASA guideline for the early management of patients with acute ischemic stroke[2]. Refinement used additional data independent of the evaluation set, and the rule set was frozen before evaluation. These rules encode the standardizable core of triage, IVT eligibility and reperfusion decision-making. To support decision-making in complex reperfusion scenarios that are not readily resolved by the rules, we constructed structured summaries of 30 primary reperfusion trials, each recording the enrollment window, study population, intervention and comparator, primary result and relevant boundary conditions. These scenarios included extended-window thrombolysis or thrombectomy, large-core or posterior-circulation infarction, combined relative contraindications, thrombolytic drug choice, and the choice between bridging and EVT alone. At final reperfusion planning, the full set of summaries is provided to the reperfusion agent as evidence context in a single pass. The agent matches each summary to the case within its own reasoning and cites a summary only when the trial’s time window, population and intervention match the case and its result directly drives the decision; cases resolved by the standard rules cite no trial. Citations are returned as a structured list giving the source, its level of evidence, and the reason the summary applies, so that each recommendation can be checked against its supporting evidence.

### 4.5 Evaluation of StrokeAgent

#### 4.5.1 Evaluation design

As shown in Extended Data Fig. 2, we evaluated StrokeAgent in four parts, all built around a staged emergency-care workflow. First, we compared its decisions with the expert reference, which was established under the same staged information boundaries and served as the reference for all comparisons. Second, we compared it with three general-purpose, tool-free LLMs (GPT-5.4, Gemini 3.1 Pro and Claude Opus 4.6) and with six resident and attending neurologists, under matched information content and timing. Third, we assessed the quality of the clinical rationale behind each decision: outputs from StrokeAgent and the three LLMs were scored under blinded review by an independent panel of raters. Fourth, we reviewed the cases in which StrokeAgent’s final decision was discordant with the expert reference to localize errors to components or to decision synthesis.

#### 4.5.2 Evaluation dataset

The evaluation set comprised 100 cases, of which 75 were real emergency cases from three tertiary hospitals in eastern China: Shanghai Sixth People’s Hospital (20 cases), Minhang Hospital of Fudan University (25 cases), and the Second Affiliated Hospital of Zhejiang University (30 cases). For these cases we collected the emergency medical records and all imaging acquired during the emergency visit. Fifteen of the real cases had no CTP imaging available, reflecting the variable availability of perfusion imaging in routine emergency practice. The remaining 25 cases came from the ISLES 2024 challenge dataset[43], which provides multimodal imaging (including CTA and 4D CTP) and a brief clinical summary; for each, expert panel constructed a complete but fictional patient record covering triage text and medical history.

This study was reviewed and approved by the Ethics Committee of Minhang Hospital, Fudan University (approval no. 2026-Approval-049-01K). The other two hospitals obtained approval from their respective institutional ethics committees (approval nos. 2026 Ethics Review Research No. 1026 and 2026-KY-289 (K)). All patient information used in the study was de-identified before analysis.

The set was deliberately enriched for clinically complex decisions. Standard cases were those meeting the unambiguous conditions of a single treatment category under the 2026 AHA/ASA guideline (a Class I or Class III recommendation), for example a typical IVT indication within the time window and without relative contraindications, or EVT alone for a proximal LVO without marked early ischemic change. Complex cases were those involving at least one of the following: an extended or uncertain time window, a relative contraindication, large-core infarction with potential thrombectomy benefit, premorbid moderate disability combined with LVO or minor but potentially disabling symptoms.

#### 4.5.3 Staged information protocol

To reproduce the order in which information arrives in the emergency department, each case was presented to StrokeAgent, the comparator groups and the expert panel in a fixed three-stage sequence. Stage 1 (emergency triage) provided the emergency admission record, bedside vital signs, fingertip blood glucose and the raw 12-lead ECG waveform. Stage 2 (initial thrombolysis decision) added the source NCCT images and the first laboratory results. Stage 3 (final reperfusion planning) added CTA and CTP. At each stage, all groups received the same underlying clinical information and imaging modalities, although the presentation format differed between groups. The expert panel and the neurologists worked in a web-based interface: clinical text, laboratory results and bedside measurements were shown as text; NCCT and CTA were viewed in an embedded DICOM viewer that allowed scrolling through the source series, with MIP and vessel reconstruction images additionally provided for CTA; and CTP was shown as post-processed perfusion maps and volumetric measurements exported from each hospital’s picture archiving and communication system (PACS). For the ISLES 2024 cases, which include perfusion parameter maps, volumes were derived using commonly used thresholds (relative cerebral blood flow *<*30% for the ischemic core and Tmax *>*6 s for hypoperfused tissue). Information from later stages was withheld from every group until the corresponding stage.

#### 4.5.4 Expert reference

The expert reference for each case was established by an expert panel of five physicians: three general neurologists and two neurointerventionalists, with 12 to 34 years of clinical experience. At each stage the panel determined the most appropriate decision using only the information released up to that stage. Decisions were informed by the 2026 AHA/ASA guideline and the panel’s clinical expertise. Every case was discussed until the panel reached consensus on both the intermediate assessments and the three decisions. Panel members worked independently of the neurologist group, of the raters who scored the reasoning outputs and of the experts who developed the clinical rules.

#### 4.5.5 Comparator groups

The three tool-free LLM baselines received the same information as StrokeAgent at each stage, with native imaging files converted into model-compatible representations: NCCT as brain-window axial slices sampled at fixed intervals (12–15 slices), CTA as axial images together with MIP reconstructions and CTP as post-processed perfusion parameter maps and quantitative measurements, the same as those provided to the human evaluators.

The neurologist group comprised six neurologists from five comprehensive stroke centers in southeastern China, with a median of 5.5 years of clinical experience (range 3–11). Before evaluation, they completed standardized training on the 2026 AHA/ASA guideline and worked through five practice cases excluded from the analysis. They viewed the materials in the interface described above and judged independently. They used no AI tools but could consult conventional clinical reference sources, and were blinded both to one another and to the expert reference. The neurologists were required to derive intermediate findings, including NIHSS, ASPECTS and LVO status before selecting a treatment.

Decision time was measured separately for each stage. For StrokeAgent, timing began when the stage input became available to the system and ended when the structured decision was returned, and included tool execution, such as generating perfusion maps and quantitative measurements from source CTP data. For neurologists, timing was recorded by the evaluation interface from opening the stage material to submission of the decision; the perfusion maps and volumetric measurements they reviewed were precomputed. End-to-end time was the sum of all applicable stage times. For each case, neurologist time was taken as the median across the six neurologists’ end-to-end times.

#### 4.5.6 Rationale quality assessment

To evaluate the quality of the clinical rationales, we adapted the framework of Ferber et al[20]. and developed a structured scoring rubric; full criteria are provided in Supplementary Information. Factual correctness assessed the accuracy of patient-specific facts and imaging findings, along with any general medical statements in the output. Completeness assessed whether the information needed to support the decision at each stage was adequately considered. Stage appropriateness evaluated whether the decision relied only on information available at that stage, without anticipating information not yet available or delaying action unnecessarily. Reasoning–decision consistency assessed whether the stated reasoning supported the decision reached. We also rated clinical safety, categorized as no, mild-to-moderate, or severe potential harm. Overall clinical utility captured the extent to which the output could support a safe, appropriate and actionable decision by a stroke physician at the relevant point in care. All dimensions except clinical safety were rated on five-point ordinal scales.

The unit of evaluation was the complete three-stage decision trajectory, including the accompanying clinical rationale, produced by one system for one case (hereafter, an output). For each output, raters were shown the case information available at each stage together with the corresponding expert reference decision.

A separate panel of six neurologists rated the outputs of StrokeAgent and the three tool-free LLMs across all 100 cases, yielding 400 case–system outputs. Review assignments followed a balanced incomplete block design: within each case, the four outputs formed six possible pairs, and each of the six raters was randomly assigned a different pair, so that each rater reviewed two outputs per case and each output received three independent ratings. Presentation order was randomized across cases, and system identity was concealed throughout.

#### 4.5.7 Error analysis

Cases in which StrokeAgent’s final decision was discordant with the expert reference were independently reviewed by the expert panel to determine the primary error mechanism. For each case, reviewers examined the source clinical record, all component and subagent outputs, and the complete decision trajectory. A component error was assigned when a specific tool or subagent output contained a clinically meaningful inaccuracy, such as a missed LVO or a substantially inaccurate ASPECTS estimate. A decision-synthesis error was assigned when the component outputs were factually accurate, with no clinically meaningful errors, but inappropriate integration of the findings, clinical rules, or supporting evidence, or a clearly flawed reasoning process, led to a discordant treatment decision. Classification disagreements were resolved by panel discussion to consensus.

### 4.6 Statistical analysis

The primary decision-performance endpoint was final clinical decision concordance, defined as the proportion of cases in which the final decision matched the expert reference. We also report stage-specific concordance at each of the three decision points and complete-pathway concordance, which required agreement at every applicable decision point. Chance-corrected agreement for the six-category final decision was summarized using unweighted Cohen’s *κ*. For the final decision, we additionally report the confusion matrix, class-specific recall, precision and F1 scores, and macro-averaged F1 as a class-balanced summary measure.

Decision concordance was compared across ten sources using logistic generalized estimating equations (GEE), with source as a fixed effect, case as the clustering unit and robust sandwich standard errors. We estimated each source’s marginal concordance and formed equally weighted contrasts on the probability scale, comparing StrokeAgent against the mean of the three LLM baselines and against the mean of the six neurologists; StrokeAgent was also compared with each LLM individually. Proportions and between-source absolute differences are reported with 95% CIs obtained from 10,000 case-level nonparametric bootstrap resamples.

Overall clinical utility was analyzed using a proportional-odds cumulative-link mixed model with a logit link, with system and rater as fixed effects and case and case–system output as random intercepts. StrokeAgent was compared with each LLM using common odds ratios and 95% CIs. The four secondary quality dimensions were analyzed using the same model. For descriptive case-level analysis, an output was classified as favorable when the median utility rating across its three raters was at least 4; paired proportions were compared using exact McNemar tests. Inter-rater agreement was quantified using ordinal Krippendorff’s *α*.

Clinical safety was analyzed separately from the quality ratings. Any potential harm and severe potential harm were defined when at least two of three raters assigned the corresponding harm category. Overall differences among the four systems were assessed using Cochran’s Q test, followed by pairwise exact McNemar tests comparing StrokeAgent with each LLM.

Decision time was summarized as median (IQR). The mean paired difference was reported with a 95% CI from 10,000 bootstrap resamples and tested using a two-sided paired t-test.

All tests were two-sided at *α* = 0.05; Holm adjustment was applied to prespecified multiple-comparison families. Statistical analyses were performed in R (version 4.6.1), with data processing and visualization in Python (version 3.9.6).

## Data Availability

The de-identified individual-level data from the participating hospitals are not publicly available because of ethical, privacy and institutional restrictions. Qualified researchers may request access by submitting a proposal to the corresponding author.

## Data availability

The data supporting the findings of this study are available within the Article and its Supplementary Information. The de-identified individual-level data from the participating hospitals are not publicly available because of ethical, privacy and institutional restrictions. Qualified researchers may request access by submitting a proposal to the corresponding author. Requests will be reviewed by the relevant institutions and ethics committees at Shanghai Minhang Hospital, Fudan University (Shanghai, China), the Second Affiliated Hospital of Zhejiang University School of Medicine (Hangzhou, China), and Shanghai Sixth People’s Hospital, Shanghai Jiao Tong University School of Medicine (Shanghai, China), to ensure compliance with applicable ethical, confidentiality and intellectual-property requirements. Access to de-identified data may be granted upon reasonable request, subject to the relevant ethics approvals, institutional requirements and data-use agreements.

## Code availability

The source code of StrokeAgent will be made publicly available on GitHub upon publication.

## Funding

This work was supported by the National Key Research and Development Program of China (grant nos. 2025YFC3608605 and 2023YFF1204800), the Key Discipline of Public Health, Minhang District, Shanghai (grant no. 2023-04 MGWXK), and the Key Discipline of Shanghai Municipal Health System (grant no. 2024ZDXK0007). The funders had no role in study design, data collection, data analysis, interpretation of the results, preparation of the manuscript or the decision to submit the manuscript for publication.

## Competing interests

The authors declare no competing interests.

## Notes

### Competing Interest Statement

The authors have declared no competing interest.

### Author Declarations

The Ethics Committee of Minhang Hospital, Fudan University gave ethical approval for this work (approval no. 2026-Approval-049-01K). The Human Ethics Committee of the Second Affiliated Hospital, Zhejiang University School of Medicine gave ethical approval for this work (approval reference: 2026 Ethics Review Research No. 1026). The Ethics Committee of Shanghai Jiao Tong University Affiliated Sixth People's Hospital gave ethical approval for this work (approval no. 2026-KY-289 (K)). All patient data were de-identified before analysis.

